# Comorbidity and Sociodemographic determinants in COVID-19 Mortality in an US Urban Healthcare System

**DOI:** 10.1101/2020.06.11.20128926

**Authors:** An-Li Wang, Xiaobo Zhong, Yasmin L Hurd

**Affiliations:** Department of Psychiatry, Icahn School of Medicine at Mount Sinai, New York, USA; Addiction Institute of Mount Sinai, Icahn School of Medicine at Mount Sinai, New York, USA; Department of Neuroscience, Icahn School of Medicine at Mount Sinai, New York, USA; Department of Population Health Science and Policy, Icahn School of Medicine at Mount Sinai, New York, USA; Tisch cancer institute, Icahn School of Medicine at Mount Sinai, New York, USA; Behavioral Health System, Mount Sinai, New York, USA

**Keywords:** COVID-19, SARS-CoV-2, comorbidities, sociodemographic determinant

## Abstract

**Background:** New York City is the US epicenter of the coronavirus disease 2019 (COVID-19) pandemic. Early international data indicated that comorbidity contributes significantly to poor prognosis and fatality in patients infected with SARS-CoV-2. It is not known to what degree medical comorbidity and sociodemographic determinants impact COVID-19 mortality in the US.

**Methods:** Evaluation of de-identified electronic health records of 7,592 COVID-19 patients confirmed by SARS-CoV-2 lab tests in New York City. Medical comorbidites and outcome of mortality, and other covariates, including clinical, sociodemographic, and medication measures were assessed by bivariate and multivariate logistic regression models.

**Results:** Of common comorbid conditions (hypertension, chronic kidney disease, chronic obstructive pulmonary disease, asthma, obesity, diabetes, HIV, cancer), when adjusted for covariates, chronic kidney disease remained significantly associated with increased odds of mortality. Patients who had more than one comorbidities, former smokers, treated with Azithromycin without Hydroxychloroquine, reside within the boroughs of Brooklyn and Queens Higher had higher odds of death.

**Conclusions:** Increasing numbers of comorbid factors increase COVID-19 mortality, but several clinical and sociodemographic factors can mitigate risk. Continued evaluation of COVID-19 in large diverse populations is important to characterize individuals at risk and improve clinical outcomes.

## Background

New York City is the epicenter in the US of the coronavirus disease 2019 (COVID-19) pandemic. As of June 11, 2020, more than 205, 405 confirmed cases and upwards of 17,300 deaths were reported across New York City’s five boroughs.[1] The disease is known to be highly contagious, approximately one of every six people infected with the virus becomes seriously ill, developing severe respiratory symptoms that can lead to death.[2] Early clinical data from China, South Korea, and Italy have indicated a high prevalence of comorbidities among COVID-19 patients suggesting that medical comorbidity contributes to poor prognosis.[3-6] These studies describe hypertension, diabetes, coronary heart disease, kidney disease, liver disease, and some cancers as associated underlying diseases.[4, 7] In a study of 1,590 COVID-19 patients in China, 25% of patients reported having at least one comorbidity, with hypertension and diabetes among the most common underlying conditions.[4] However, interpretation of these results was limited to a highly racially homogeneous population, relatively small sample size, and limited consideration of covariates. Exploration of the role that medical comorbidity and demographic determinants play in COVID-19 mortality among patients in the US has thus far been lacking.[8, 9] Study of the impact of medical comorbidity on COVID-19 mortality in New York City, with its large, highly heterogeneous population, can offer important insights into the disease.

Our study presents a large clinical dataset on the rates of fatality and associated comorbidities in all confirmed COVID-19 patients within the Mount Sinai Health System (MSHS) in New York City. MSHS is the largest integrated healthcare delivery system in the New York City metropolitan region serving 3,499,000 million outpatients and 152,520 inpatients annually and, as such, serves as a proxy for the City’s population. We studied the impact of comorbidity on COVID-19 mortality adjusted for other relevant clinical characteristics, and assessed potential sociodemographic impacts on clinical outcomes.

## Methods

### Participants

Seven thousand five hundred and ninety-two (7,592) confirmed COVID-19 patients as April 15, 2020 within MSHS in New York City (NYC) were included in this retrospective study. MSHS comprises eight hospitals and more than 400 ambulatory clinics throughout the five boroughs of the City (Brooklyn, the Bronx, Manhattan, Queens, and Staten Island). Patients were included in the study if their laboratory results in electronic health records (EHR) stated SARS-CoV-2 positivity.

### Procedures

All study data were automatically deidentified and retrieved from the EHR. To protect privacy and comply with the Health Insurance Portability and Accountability Act (HIPAA), the following steps were taken to de-identify this data set: (1) Patient medical record number and Encounter ID were replaced with surrogate identifiers; (2) Patient date of birth was converted to age at the time of encounter; (3) All dates were converted to calculated elapsed days since the encounter date to preserve the temporal relationship among events; (4) Zip codes were truncated to the first three digits; (5) All other identifiers as specified in HIPAA’s Privacy Rule were masked or removed.

Diagnosis of comorbidities, including hypertension, diabetes, chronic kidney disease, chronic obstructive pulmonary disease (COPD), obesity, cancer, asthma and human immunodeficiency virus (HIV) were indicated as “Active” on the “patient’s problem list.” Mortality was indicated by a “deceased” marker. Ethnicity was reported as Non-Hispanic, Hispanic or Unknown. Race was classified as white, African American, Asian, other (i.e. native Hawaii, American Indian, and unspecific) and unknown (i.e. missing data). Residential area defined by the first 3 digits of the patients’ zip code was categorized for all 5 NYC boroughs, outside NYC but within NY state, and non-NY. Smoking status was described for tobacco usage as yes (i.e. current smokier), quit (i.e. former smoker), never (i.e. never smoker). If there was no value, data was treated as missing. Patient’s hospitalization status was indicated as inpatient, outpatient, emergency department, and unspecified. Vital signs were recorded during the first initial clinical encounter: a) Temperature referred to patient’s first recorded temperature. b) Temperature max referred to highest recorded temperature. c) Systolic BP referred to the first systolic blood pressure reading. d) Diastolic BP referred to the first diastolic blood pressure reading. e) O2 saturation referred to the first recorded oxygen saturation. f) O2 saturation min referred to the patient’s lowest recorded oxygen saturation. g) Respiratory rate referred to the first recorded respiratory rate. h) Heart rate referred to the first recorded heart rate. Six COVID-19 related medications were administered within MSHS, including Hydroxychloroquine, Azithromycin, Tocilizumab, Sarilumab, Anakinra, and Remdesivir. However, with the exception of Hydroxychloroquine and Azithromycin, only a few patients were administered other medications. We included only Hydroxychloroquine and Azithromycin for analysis.

### Statistical analysis

Descriptive statistics of demographic, patient, and treatment-related variables were quantified by frequencies (with percentage) for categorical variables and the means (with standard deviation) for continuous variables for the entire study population. We compared characteristics between patients with and without comorbidities using a chi-square test for categorical variables and a t-test for continuous variables. Mortality was defined as a binary variable, with yes indicating a patient died as of April 15, 2020. Associations between the outcome of mortality and comorbidities, as well as other covariates, were assessed by bivariate and multivariate logistic regression models. For each patient, we created a series of binary variables, each of which indicates the presence or absence of a type of comorbidity (e.g., hypertension, diabetes, etc.). We then created a variable summarizing the total number of comorbidities observed in each patient. All these variables were included in logistical regression. We first conducted bivariate analysis by building a series of logistic regression models, each of which examined the association between the outcome of mortality and every single predictor. Those predictors with significant associations with mortality at 0.05 level were included in the multivariate analysis. A backward selection algorithm was applied for variable selection in building the final multivariable logistic regression; any covariate with a P-value less than 0.15 was excluded from the final model. Interactions between risk factors selected in the final multivariable model were tested, and any interaction term with P-value less than 0.05 was included in the final model. Patients with missing values under a categorical variable (e.g., smoking status) were grouped as a subcategory termed “unknown” in analysis, and the corresponding odds ratios were reported with 95% confidence intervals. Patients with missing values in vital signal (e.g., Respiratory rate) were excluded in the analysis. All statistical tests and confidence intervals were two-sided. All analyses were conducted using SAS statistical software (SAS Institute. 2019).

### Patient and public involvement

This was a retrospective case series study and no patients were involved in the study design, setting the research questions, or the outcome measures directly. No patients were asked to advise on interpretation or writing up of results.

## Results

### Characteristics of patients with comorbidity

We studied a total of 7,592 confirmed COVID-19 patients who were registered within the Mount Sinai Health System in New York City. 2,879 (38%) patients had at least one comorbidity (Figure 1A), among which 41%, 33%, 18%, 6%, and 2% with 1, 2, 3, 4, and ≥5 comorbidities. Hypertension (N [%];1923 [25.3%]) and diabetes (1374 [18.1%]) were the two most prevalent comorbidities, followed by chronic kidney disease (646 [8.5%]), obesity (530 [7.0%]), cancer (506 [6.7%]), asthma (344 [4.5%]), chronic obstructive pulmonary disease (COPD; 191 [2.5%]), and HIV (126 [1.7%]) (Figure 1B). Female (40%) and Hispanic (47%) were more likely to have comorbidity than male (36%) and non-Hispanic (38%). African Americans (44%) had the highest with the lowest proportions of comorbidity with Asian (40%) and those where race was not identified (35%) (Table 1). In adults, the proportion of patients with comorbidity increased as age increased. However, the proportion of patients with comorbidity in the group under 18 years of age (24%) was higher than that of patients 18-39 years of age (15%). The proportion of those with comorbidity was significantly higher among former smokers (70%) compared to the proportions in patients who never smoked (42%) and current smokers (45%). Across NYC boroughs, residents in Manhattan and Brooklyn had the highest (43%) and lowest (27%) proportion of comorbidity, respectively. Compared to patients without comorbidity, those with comorbidity had significantly higher maximum temperatures, systolic BP, and respiratory rate, but significantly lower diastolic BP and minimum oxygen saturation, for the initial clinical measurements. Compared patients without comorbidity, more patients with comorbidity received at least one medication of Hydroxychloroquine and Azithromycin. 38%, 9%, and 11% of COVID-19 patients with at least one comorbidity received treatments of Hydroxychloroquine combined with Azithromycin, Hydroxychloroquine without Azithromycin, and Azithromycin without Hydroxychloroquine. Percentages for those without comorbidity were 26%, 6%, and 6%, respectively (Figure 2).

**Table 1:**
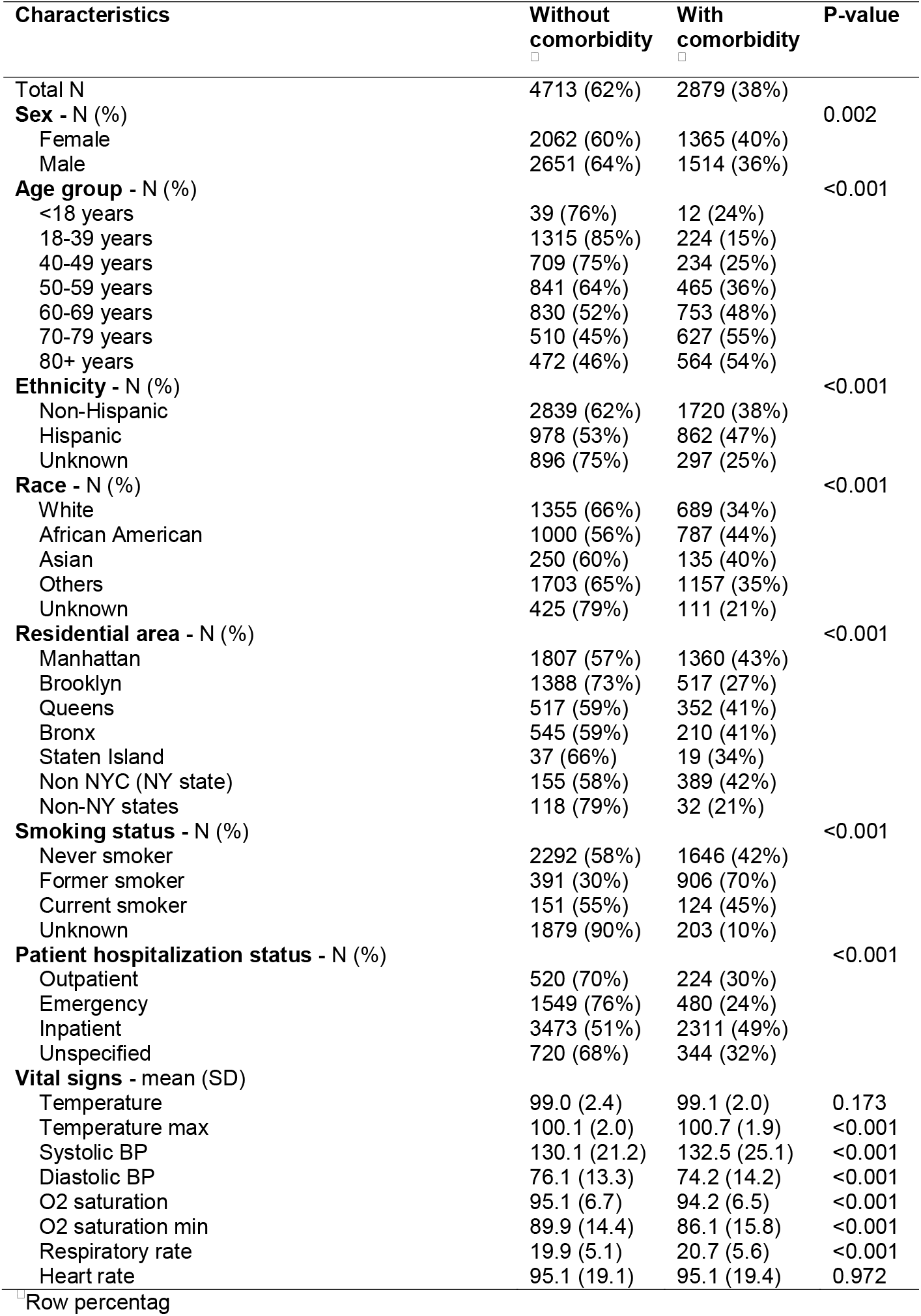
Demographic and clinical characteristics.

**Figure 1:**
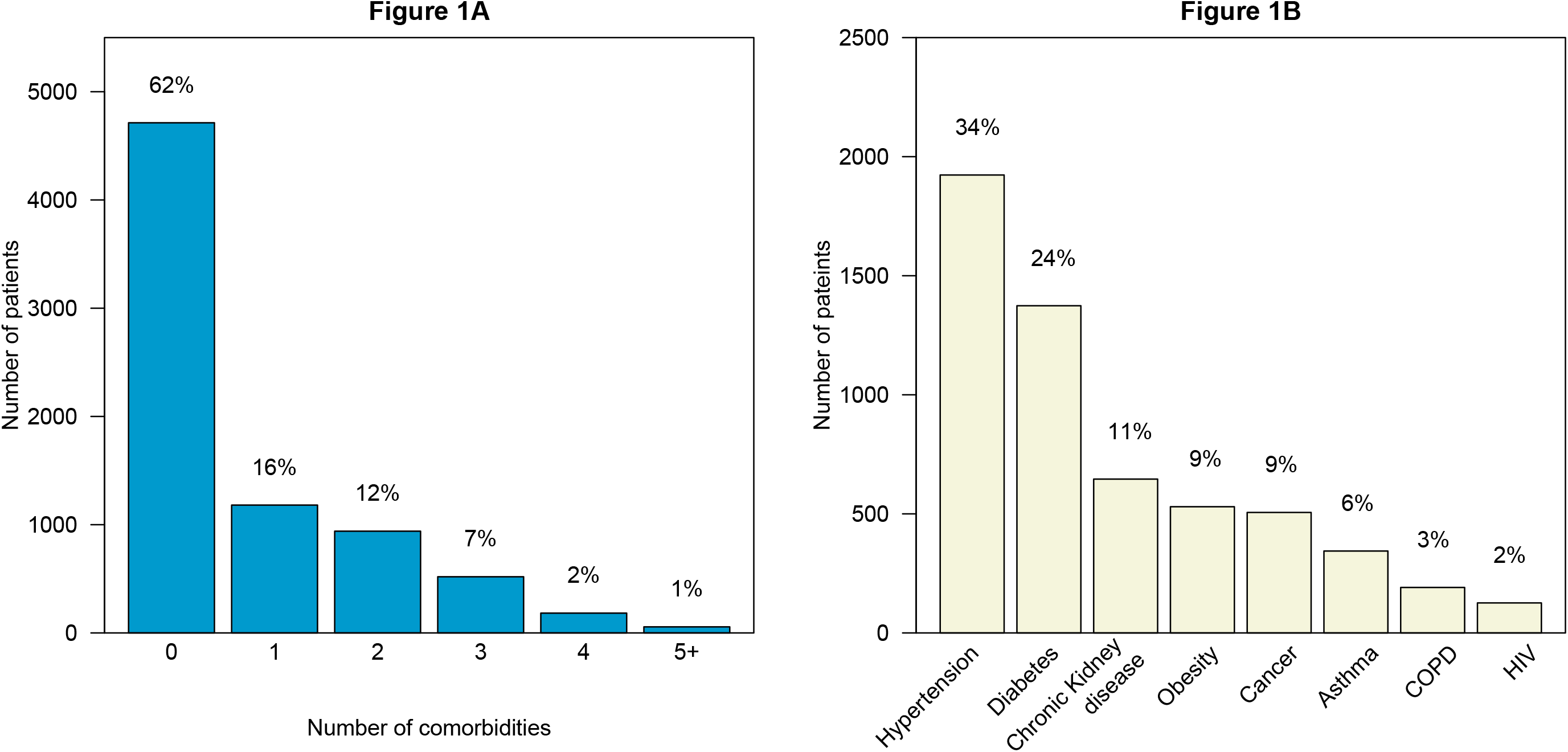
(A) Percentage and frequencies of medical comorbidities (B) Percentage and frequency of each type of medical comorbidity

**Figure 2.**
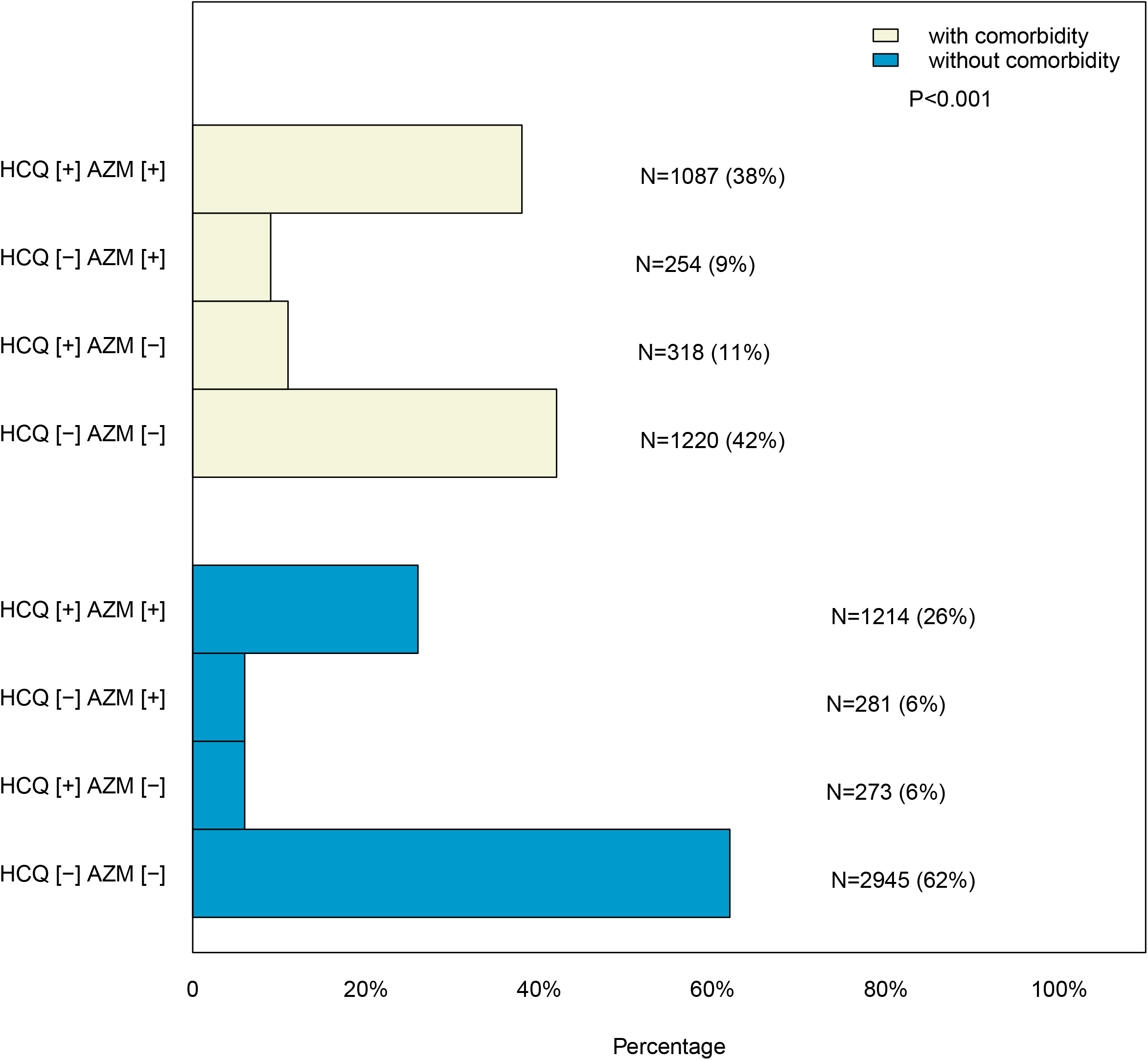
The administration of Hydroxychloroquine (HCQ) and Azithromycin (AZM) between patients with and without comorbidity in Mount Sinai Health System.

### COVID-19 mortality and bivariate analysis

Table 2 shows case fatality rates across different subgroups for different risk factors and results of bivariate analysis. Bivariate logistic regression analysis showed that COPD (OR: 2.45 [95% CI: 1.73-3.47], P<0.001), hypertension (OR: 2.33 [2.01-2.71], P<0.001), chronic kidney disase (OR: 2.38 [1.94-2.93], P<0.001), diabetes (OR: 1.97 [1.67-2.32], P<0.001) and cancer (OR: 1.29 [0.99-1.69], P=0.059) were associated with an increased odds ratio of mortality. The corresponding case fatality rate by the type of comorbidity is shown in Figure 3. The number of comorbidities is important as it has significant impact on the outcome of mortality; a greater number of comorbidities was associated with a higher case fatality rate (1-3 comorbidities: OR: 2.10 [1.81-2.44], P<0.001; 4+ comorbidities: OR: 2.68 [1.90-3.76], P<0.001) (Figure 4). Other risk factors associated with a higher rate of fatality included sex (male) (OR: 1.25 [95% CI: 1.08-1.44], P=0.003), former smokers (OR: 1.78 [1.49-2.13], P<0.001), inpatient hospitalization (OR: 36.95 [15.28-86.36], P<0.001), emergency department (OR: 4.97 [1.99-12.38], P<0.001), administration of Azithromycin but no Hydroxychloroquine (OR: 7.18 [5.57-9.25], P<0.001), combined Azithromycin and Hydroxychloroquine (OR: 5.55 [4.60-6.68], P<0.001), or Hydroxychloroquine but no Azithromycin (OR: 4.90 [3.77-6.39], P<0.001), as well as residency of the patient (within NY state but outside of New York City (OR: 2.10 [1.70-2.58], P<0.001); Brooklyn (OR: 1.49 [1.25-1.79], P<0.001); and Queens (OR: 1.22 [0.95-1.56], P<0.001). Older age was also significantly associated with increased odds of mortality. Specifically, compared to COVID-19 patients under the age of 40, OR: 5.75 [2.82, 11.72]; 50-59 years: OR: 10.31 [5.32-19.98, P<0.001]; 60-69 years: 18.63 [9.81-35.11], P<0.001; 70-79 years: 37.27 [19.67-70.61], P<0.001, and 80+ years: 80.93 [37.57-133.93], P<0.001). Odds of death for Hispanic patients with COVID-19 were significantly lower than for non-Hispanic patients (OR: 0.81 [0.68, 0.97], P=0.022).

**Table 2:** Case fatality rates by risk factors and unadjusted odds ratios of mortality with 95% confidence interval.

| Risk factors | Deceased |  | unadjusted Odds ratio (95% CI) | P-value |
| --- | --- | --- | --- | --- |
|  | No | Yes |  |  |
| **Sex** |  |  |  |  |
| Female | 3093 (90%) | 334 (10%) | reference group |  |
| Male | 3671 (88%) | 494 (12%) | 1.25 (1.08, 1.44) | 0.003 |
| **Age group** |  |  |  |  |
| <40 years | 1580 (99%) | 10 (<1%) | reference group |  |
| 49-49 years | 907 (96%) | 33 (4%) | 5.75 (2.82, 11.72) | <0.001 |
| 50-59 years | 1226 (94%) | 80 (6%) | 10.31 (5.32, 19.98) | <0.001 |
| 60-69 years | 1416 (89%) | 167 (11%) | 18.63 (9.81, 35.41) | <0.001 |
| 70-79 years | 920 (81%) | 217 (19%) | 37.27 (19.67, 70.61) | <0.001 |
| 80+ years | 715 (69%) | 321 (31%) | 70.93 (37.57, 133.93) | <0.001 |
| **Ethnicity** |  |  |  |  |
| Non-Hispanic | 4011 (88%) | 548 (12%) | reference group |  |
| Hispanic | 1656 (90%) | 184 (10%) | 0.81 (0.68, 0.97) | 0.022 |
| Unknown | 1097 (92%) | 96 (8%) | 0.64 (0.51, 0.80) | <0.001 |
| **Race** |  |  |  |  |
| White | 1788 (88%) | 236 (12%) | reference group |  |
| African American | 1580 (88%) | 207 (12%) | 0.99 (0.81, 1.21) | 0.941 |
| Asian | 348 (90%) | 37 (10%) | 0.81 (0.56, 1.16) | 0.246 |
| Others | 2546 (89%) | 314 (11%) | 0.93 (0.78, 1.12) | 0.458 |
| Unknown | 502 (94%) | 34 (6%) | 0.51 (0.35, 0.75) | <0.001 |
| **Residential area** |  |  |  |  |
| Manhattan | 2877 (91%) | 290 (9%) | reference group |  |
| Brooklyn | 1656 (87%) | 249 (13%) | 1.49 (1.25, 1.79) | <0.001 |
| Queens | 774 (89%) | 95 (11%) | 1.22 (0.95, 1.56) | <0.001 |
| Bronx | 488 (96%) | 23 (5%) | 0.47 (0.30, 0.72) | 0.115 |
| Staten Island | 51 (91%) | 5 (9%) | 0.97 (0.39, 2.46) | 0.953 |
| Non NYC (NY state) | 772 (83%) | 163 (17%) | 2.10 (1.70, 2.58) | <0.001 |
| Non-NY state | 147 (98%) | 3 (2%) | 0.20 (0.06, 0.64) | 0.007 |
| **Smoking status** |  |  |  |  |
| Never smoker | 3544 (90%) | 394 (10%) | reference group |  |
| Former smoker | 1083 (84%) | 214 (17%) | 1.78 (1.49, 2.13) | <0.001 |
| Current smoker | 247 (90%) | 28 (10%) | 1.02 (0.68, 1.53) | 0.925 |
| Unknown | 1890 (91%) | 192 (9%) | 0.91 (0.76, 1.10) | 0.330 |
| **Patient hospitalization status** |  |  |  |  |
| Outpatient | 739 (99%) | 5 (<1%) | reference group |  |
| Emergency | 1963 (97%) | 66 (3%) | 4.97 (1.99, 12.38) | <0.001 |
| Inpatient | 3004 (80%) | 817 (14%) | 36.95 (15.28, 89.36) | <0.001 |
| **Vital signs** |  |  |  |  |
| Temperature | - | - | 0.973 (0.948, 0.999) | 0.045 |
| Temperature max | - | - | 1.262 (1.214, 1.311) | <0.001 |
| Systolic BP | - | - | 0.995 (0.991, 0.998) | 0.001 |
| Diastolic BP | - | - | 0.976 (0.971, 0.982) | <0.001 |
| O2 saturation | - | - | 0.923 (0.914, 0.933) | <0.001 |
| O2 saturation min | - | - | 0.926 (0.920, 0.932) | <0.001 |
| Respiratory rate | - | - | 1.068 (1.055, 1.080) | <0.001 |
| Heart rate | - | - | 1.003 (0.999, 1.006) | 0.170 |
| **Administered med at MSHS** |  |  |  |  |
| HCQ [-] AZM [-] | 3997 (96%) | 168 (4%) | reference group |  |
| HCQ [+] AZM [-] | 490 (83%) | 101 (17%) | 4.90 (3.77, 6.39) | <0.001 |
| HCQ [-] AZM [+] | 411 (77%) | 124 (23%) | 7.18 (5.57, 9.25) | <0.001 |
| HCQ [+] AZM [+] | 1866 (81%) | 435 (19%) | 5.55 (4.60, 6.68) | <0.001 |

**Figure 3:**
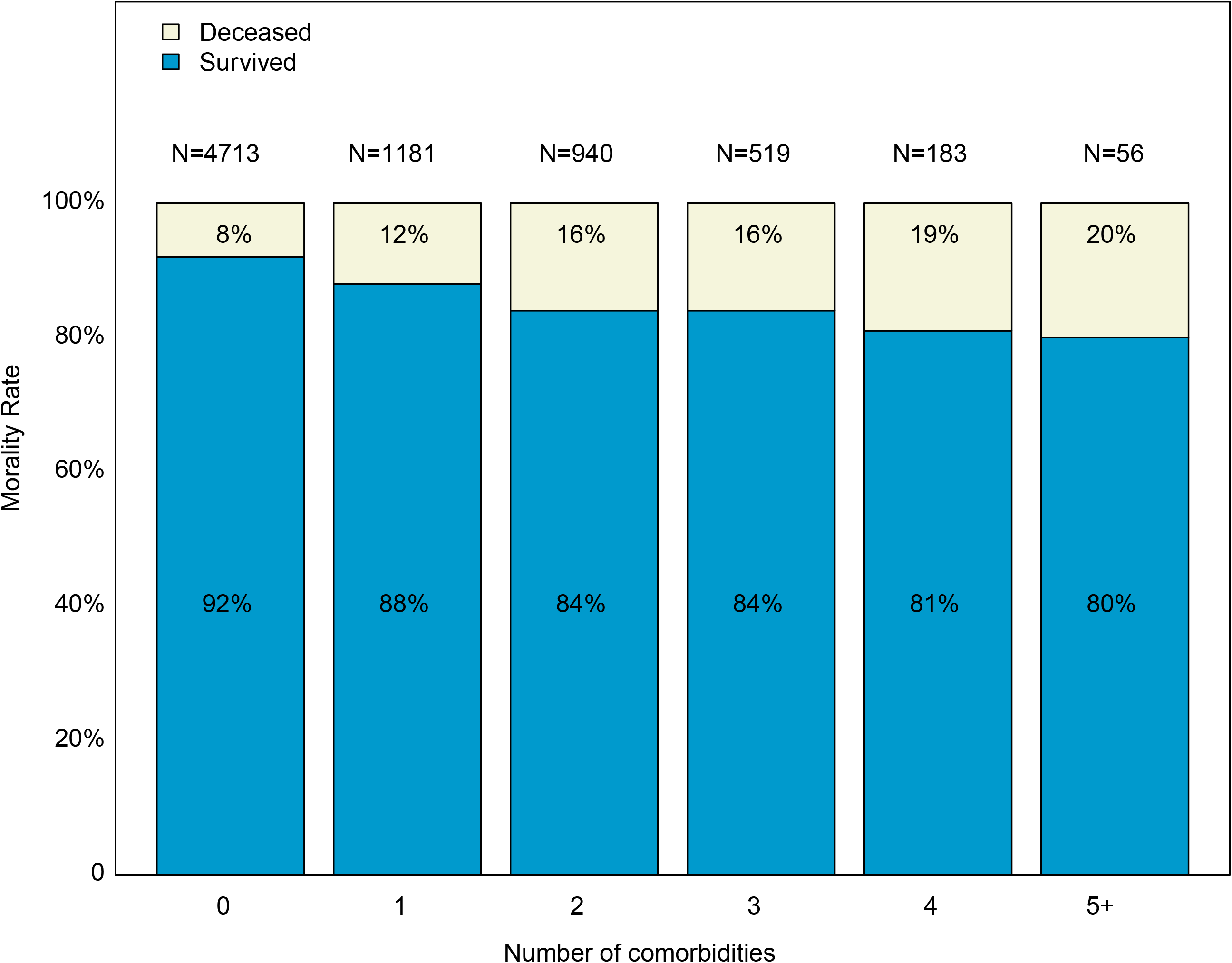
Case fatality rate of COVID-19 patients with or without medical comorbidity.

**Figure 4:**
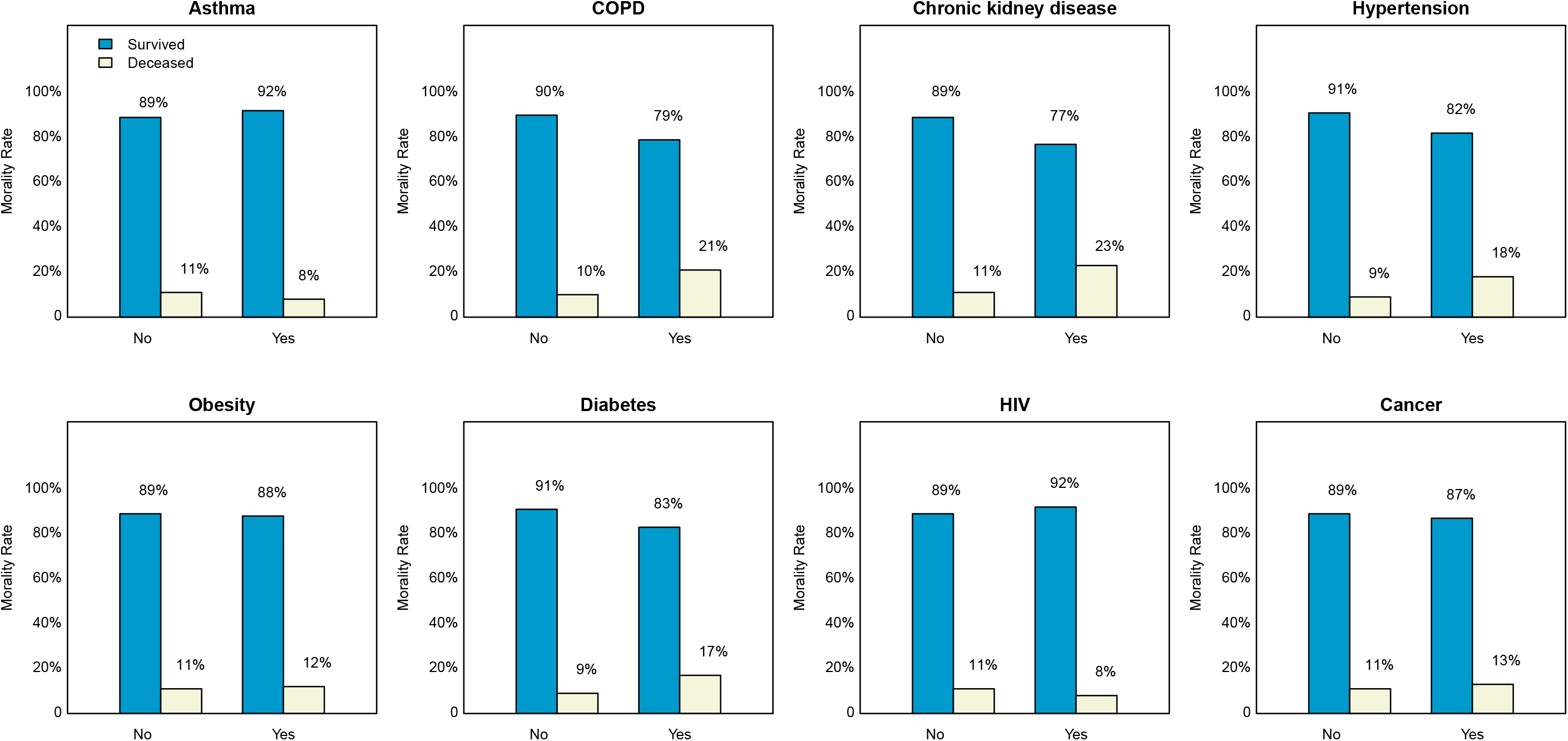
Case fatality rate of COVID-19 patients based on the number of comorbidities.

### Adjusted multivariable evaluation of comorbidity and COVID-19 mortality

Table 3 provides adjusted estimated ORs (95% CIs) and P-values of the final multivariable model. Adjusted for covariates, the multivariable logistic regression model revealed that chronic kidney disease (OR: 1.36 [1.01-1.83], P=0.042) was statistically significantly associated with increased odds of mortality. Asthma was marginally significantly associated with lower odds of mortality (OR: 0.63 [0.38-1.04], P=0.073). Compared to those without comorbidity, COVID-19 patients with no more than three comorbidities had significantly higher odds of mortality (OR: 1.27 [1.02-1.58, P=0.029]). Although the odds of mortality tended to be higher in COVID-19 patients with more than 3 comorbidities (OR: 1.71 [0.96-3.04], P=0.067), this trend was not statistically significant at 0.05 level. Higher odds of mortality were also significantly associated with higher maximum temperature (OR: 1.172 [1.115-1.231], P=0.002), faster respiratory rate (OR: 1.025 [1.010-1.041], P=0.001), former smokers (OR: 1.27 [1.01-1.60], P=0.041), residency (in NY state but outside of New York City (OR: 2.12 [1.63-2.75], P<0.001); Brooklyn (OR: 2.33 [1.85-2.95], P<0.001); Queens (OR: 1.42 [1.05-1.94], P=0.023)), and administration of Azithromycin without Hydroxychloroquine (OR: 1.57 [1.14-2.16], P=0.006). Patients with higher systolic BP (OR: 0.994 [0.991-0.998], P=0.002) and higher minimum oxygen saturation (OR: 0.942 [0.936-0.948], P<0.001) during the first clinical evaluation has lower odds of death. The likelihood of death for COVID-19 patients significantly increased with age (40-49 years vs. <40 years OR: 3.48 [1.48-8.71], P=0.004). Odds of death nearly doubled for every 10 years’ increase among patients older than 50 (50-59 years: OR: 5.53 [2.51-12.19, P<0.001]; 60-69 years: 8.44 [3.90-18.26], P<0.001; 70-79 years: 18.29 [8.46-39.51], P<0.001, and 80+ years: 33.77 [15.66-72.84], P<0.001). No significant interaction was found between comorbidity and other covariates.

**Table 3:** Adjusted multivariate evaluation of comorbidity and COVID-19 mortality. Adjusted odds ratio of mortality with 95% confidence interval.

| Risk factors | Adjusted Odds ratio (95% CI) | P-value |
| --- | --- | --- |
| **Age group** |  |  |
| <40 years | reference group |  |
| 49-49 years | 3.48 (1.48, 8.17) | 0.004 |
| 50-59 years | 5.53 (2.51, 12.19) | <0.001 |
| 60-69 years | 8.44 (3.90, 18.26) | <0.001 |
| 70-79 years | 18.29 (8.46, 39.51) | <0.001 |
| 80+ years | 33.77 (15.66, 72.84) | <0.001 |
| **Residential area** |  |  |
| Manhattan | reference group |  |
| Brooklyn | 2.33 (1.85, 2.95) | <0.001 |
| Queens | 1.43 (1.05, 1.94) | 0.023 |
| Bronx | 0.72 (0.43, 1.21) | 0.216 |
| Staten Island | 2.49 (0.86, 7.16) | 0.092 |
| Non NYC (NY state) | 2.12 (1.63, 2.75) | <0.001 |
| Non-NY state | 0.49 (0.11, 2.16) | 0.349 |
| **Smoking status** |  |  |
| Never smoke | reference group |  |
| Former smoker | 1.27 (1.01, 1.60) | 0.041 |
| Current smoke | 1.44 (0.87, 2.37) | 0.156 |
| Unknown | 1.87 (0.94, 1.50) | 0.154 |
| **Vital signs** |  |  |
| Temperature max | 1.172 (1.115, 1.231) | <0.001 |
| Systolic BP | 0.994 (0.991, 0.998) | 0.002 |
| O2 saturation min | 0.942 (0.936, 0.948) | <0.001 |
| Respiratory rate | 1.025 (1.010, 1.041) | 0.001 |
| **Administered med at MSHS\*** |  |  |
| HCQ [-] AZM [-] | reference group |  |
| HCQ [+] AZM [-] | 0.96 (0.69, 1.34) | 0.822 |
| HCQ [-] AZM [+] | 1.57 (1.14, 2.16) | 0.006 |
| HCQ [+] AZM [+] | 0.94 (0.73, 1.21) | 0.631 |
| **Medical Comorbidity** |  |  |
| Asthma |  |  |
| No | reference group |  |
| Yes | 0.63 (0.38, 1.04) | 0.073 |
| Chronic kidney disease |  |  |
| No | reference group |  |
| Yes | 1.36 (1.01, 1.83) | 0.042 |
| **Number of comorbidity** |  |  |
| 0 | reference group |  |
| 1-3 | 1.27 (1.02, 1.58) | 0.029 |
| 4+ | 1.71 (0.96, 3.04) | 0.067 |

\* HCQ, Hydroxychloroquine; AZM, Azithromycin.

## Discussion

In line with available clinical data from China, Italy, and elsewhere in the US,[3, 6, 10] risk factors for increased COVID-19 mortality related to older age and those with more severe symptoms. The rate of fatality was almost double among patients with these risk factors than those without. Although hypertension and diabetes were the most common comorbidities among COVID-19 patients and were each independently associated with the increased rate of death, chronic kidney disease contributed significantly to mortality after correcting for other confounders, including age, sex, race, ethnicity, smoking status, vitals signs, geographical residence, and medications.

With regard to other comorbid respiratory disorders and in contrast to the CDC’s list of higher risk populations,[11] in the current study, asthma was a protective factor. Indeed, Jackson and colleagues suggested that asthma may not be a risk factor because of reduced angiotensin-converting enzyme-2 (ACE2) gene expression in airway cells of asthma patients that would be expected to decrease the severity of SARS-CoV-2 infection which uses ACE2 as its cellular receptor} instead [12]. The potential protective aspect of asthma in regard to mortality risk will need to be evaluated in a larger population as the number of subjects with asthma in the current cohort was small.

In this study, past smoking significantly increased risk of mortality even when adjusted for potential confounds. Interestingly, two studies in China examined the relationship between smoking and severity of COVID-19 and found no significant correlation between smoking and rates of death.[10, 13] These results, however, may reflect their relatively small sample sizes, the narrowly defined population (exclusively Asian patients) and only current smokers. In our larger, more heterogenous confirmed COVID-19 cohort in New York City, we found a strong correlation between former smoking and COVID-19 mortality. This correlation persisted even after adjusting for confounding factors (e.g.age, sex, geographical zip code and medical comorbidity).

Social determinants of health are important considerations for comorbidities and health outcomes. Here, the zip code associated with patients’ residence was used as a partial surrogate measure of socioeconomic status (SES) given the heterogenous boroughs in NYC. Despite a greater number of comordities, those living in Manhattan had lower case fatality rates than patients residing in Brooklyn and Queens. Whether this correlation can be attributed to SES or other factors needs to be explored since we did not have information regarding attributes such as income level or education. There was, however, no significant difference in mortality risk with regard to race or ethnicity when covariates were considered.

Hydroxychloroquine, an anti-malaria medication, has been widely speculated as a potential treatment for COVID-19[14, 15] and is frequently given in combination with Azithromycin, an antibiotic medication speculated to improve COVID-19 due to its potential antiviral properties.[5] Both medications were co-administered in the current population and a small clinical trial has shown encouraging results.[16] Administration of Azithromycin and Hydroxychloroquine was associated with increased odds of mortality among COVID-19 patients with comorbidity, but only Azithromycin administration without Hydroxychloroquine was significant after adjusting for confounding factors. Recent warnings have been released by the FDA about the use of Hydroxychloroquine and more studies are critical to determine its risk/benefit profile for COVID-19.[17] However, by controling different covariates in its statistical model, a recent study reported that treatment with Azithromycin, Hydroxychloroquine, or both was not differently impact on COVID-19 mortality.[18] The discrepancy of findings emphasizes the importance of scrutinizing statistical methods when comparing results across studies, and the need for randomized control clinical trials.

The current study has several limitations especially as related to a retroactive evaluation of de-identified electronic health records. We were unable to explore in-depth individual medical cases or to access to all aspects of clinical care. Data regarding vital sign measures were limited to the initial clinical encounter and not available for the course of hospitalization for inpatients. Although the cohort studied represents a heterogenous population, whether the results generalize to other large metropolitan communities needs to be addressed as more data becomes available across the US and other countries. Another limitation is that SES cannot be accurately determined only by zip code, and caution should be undertaken in interpreting the geographic data.

In summary, the current findings confirm the critical nature of comorbid factors, as well as the increasing number of comorbidities (hypertension, chronic kidney disease, COPD, and diabetes) to mortality risk and also suggest that geographical location, which potentially relates to SES within a large metropolitan area, may increase mortality. Although females, African Americans, and Hispanics often have a greater number of comorbidities, these groups were not associated with increased mortality risk in the COVID-19 population after covarying for other factors. Continued analysis of larger populations locally and globally is essential as the pandemic evolves. Moreover, it will be important to monitor the long-term health impact of those who recovered from COVID-19 infection on the course of their comorbid health conditions.

## Data Availability

The data is available upon request.

